# Blood biomarker profiles in young-onset neurocognitive disorders: a cohort study

**DOI:** 10.1101/2024.06.30.24309746

**Authors:** Oneil G. Bhalala, Jessica Beamish, Dhamidhu Eratne, Patrick Summerell, Tenielle Porter, Simon M. Laws, Matthew JY Kang, Aamira J. Huq, Wei-Hsuan Chiu, Claire Cadwallader, Mark Walterfang, Sarah Farrand, Andrew H. Evans, Wendy Kelso, Leonid Churilov, Rosie Watson, Nawaf Yassi, Dennis Velakoulis, Samantha M. Loi

## Abstract

**INTRODUCTION:** Young-onset neurocognitive symptoms result from a heterogeneous group of neurological and psychiatric disorders which present a diagnostic challenge. To identify such factors, we analysed the BeYOND (Biomarkers in Younger-Onset Neurocognitive Disorders) cohort, a study of individuals less than 65 years old presenting with neurocognitive symptoms for a diagnosis and who have undergone cognitive and biomarker analyses.

**METHODS:** Sixty-five participants (median age at assessment of 56 years, 45% female) were recruited during their index presentation to the Royal Melbourne Hospital Neuropsychiatry Centre, a tertiary specialist service in Melbourne, Australia, and categorised as either early-onset Alzheimer’s disease (EOAD, n=18), non-AD neurodegeneration (nAD-ND, n=23) or primary psychiatric disorders (PPD, n=24). Levels of neurofilament light chain, glial fibrillary acidic protein and phosphorylated-tau 181, apolipoprotein E genotype and late-onset AD polygenic risk scores were determined. Information-theoretic model selection identified discriminatory factors.

**RESULTS:** Neurofilament light chain, glial fibrillary acidic protein and phosphorylated-tau 181 levels were elevated in EOAD compared to other diagnostic categories. A multi-omic model selection identified that a combination of cognitive and blood biomarkers, but not the polygenic risk score, discriminated between EOAD and PPD (AUC≥0.975, 95% CI: 0.825-1.000). Phosphorylated-tau 181 alone significantly discriminated between EOAD and nAD-ND causes (AUC=0.950, 95% CI: 0.877-1.00).

**DISCUSSION:** Discriminating between EOAD, nAD-ND and PPD causes of young-onset neurocognitive symptoms is possible by combining cognitive profiles with blood biomarkers. These results support utilising blood biomarkers for the work-up of young-onset neurocognitive symptoms and highlight the need for the development of a young-onset AD-specific polygenic risk score.

## Plain Language Summary

### TITLE

Use of blood tests to determine the cause of memory and thinking symptoms in young people.

### ABSTRACT

It is difficult to determine the cause of memory and thinking symptoms when they occur in young people who are under the age of 65 years. Their symptoms can be due to many different causes like psychiatric conditions, such as depression, or dementia, such as Alzheimer’s disease. It is difficult to make an accurate diagnosis because symptoms that young people experience are often very similar if the symptoms are due to psychiatric conditions or dementia. This often leads to an incorrect diagnosis and a delay in appropriate treatments.

In dementia, proteins are released by brain cells at very low levels into the blood. With new technology, blood tests can now detect some of these proteins. Studies have shown that using a blood test to measure protein can help identify people who are older than 65 years and have dementia or another cause for their thinking and memory symptoms. However, it is not known if similar blood tests can be used in people younger than 65 years.

In this study, we recruited 65 young people who were experiencing memory and thinking symptoms and performed blood tests and thinking tests. Eighteen people were diagnosed with Alzheimer’s disease, 23 were diagnosed with a cause of dementia other than Alzheimer’s disease and 24 had a primary psychiatric disorder. We found that on average, people with Alzheimer’s disease had higher blood levels of three proteins – neurofilament light chain, glial fibrillary acidic protein and phosphorylated tau-181, than those people with another diagnosis. We found that a person’s performance on thinking tests, combined with their blood levels of phosphorylated tau-181 and neurofilament light chain, may help predict if that person has Alzheimer’s disease or another condition. This study demonstrates the need for more research in diagnosing the cause of memory and thinking problems in young people.

## Background

Young-onset dementia, often defined as dementia diagnosed before the age of 65 years(van de Veen et al., 2021), is comprised of a heterogenous group of disorders that accounts for approximately 5% of all cases of dementia and have a prevalence of 119 per 100,000 individuals worldwide.(Hendriks et al., 2021; Loi et al., 2023) Diagnosing young-onset dementia is challenging due to the atypical symptoms and cognitive profiles that can overlap with other causes of neurocognitive disorders in this age group, such as primary psychiatric disorders (PPD). Nearly one-third of individuals are misdiagnosed with PPD prior to the identification of a neurodegenerative cause and nearly 40% of individuals with young-onset neurocognitive symptoms have their initial diagnosis revised during follow-up, reflecting low diagnostic certainty with standard clinical work-up.(Woolley et al., 2011; Tsoukra et al., 2022) Establishing a timely and accurate diagnosis is important to facilitate appropriate management and understand prognosis.

Blood biomarkers are emerging as powerful predictors of neurodegenerative conditions. Improvements in immunoassay technology, such as single molecule array (Simoa), have yielded ultra-sensitive detection of protein biomarkers in blood, where concentrations are often in the pico- and femtomolar ranges.(Mankhong et al., 2022) Neurofilament light chain (NfL) and glial fibrillary acidic protein (GFAP) are detected in blood samples from a variety of neurodegenerative conditions including late-onset Alzheimer’s disease (LOAD), Lewy body disease and frontotemporal dementia.(Eratne et al., 2024; Hansson et al., 2023; Bacioglu et al., 2016) Furthermore, hyperphosphorylated species of tau, such as those phosphorylated at threonine 181 (p-tau181), have demonstrated high specificity in distinguishing LOAD from other neurodegenerative conditions.(Janelidze et al., 2020)

While different protein blood biomarkers reflect dynamic changes occurring in the setting of neurodegeneration, they vary in their ability to predict onset of disease prior to symptom onset. Contrastingly, genetics can be considered as capturing an individual’s static risk of developing neurodegeneration.(Bhalala et al., 2024) Genetic variants have been identified using genome-wide association studies (GWAS) for conditions such as LOAD, Lewy body disease and Parkinson’s disease.(Kunkle et al., 2019; Chia et al., 2021; Bellenguez et al., 2022; Kim et al., 2024) Polygenic risk scores (PRS) have been derived from GWAS findings to calculate an individual’s risk of developing a particular neurodegenerative condition. For example, those with the highest 10% PRS values have nearly a 1.9-fold increase in the risk of LOAD compared to those in the lowest 10%, and this effect is additive to age and the apolipoprotein E (*APOE*) status, with the latter being the gene most strongly associated with LOAD.(Bellenguez et al., 2022)

Leveraging multi-omic data by combining the static risk captured by genetic analyses such as the PRS, the dynamic risk identified by protein biomarkers, and standard clinical assessments can improve diagnostic accuracy for late-onset neurodegenerative conditions such as LOAD.(Palmqvist et al., 2021; Ramanan et al., 2023; Bhalala et al., 2024) However, there is a paucity of data on how such an approach applies to diagnosing individuals with young-onset neurocognitive symptoms. Identifying clinical variables and blood biomarkers that are useful in discriminating between young-onset dementia causes may improve diagnostic accuracy and timeliness.

In this study, we sought to address these questions by analysing the Biomarkers in Younger-Onset Neurocognitive Disorders (BeYOND) cohort(Loi, 2020), a clinical cohort of individuals experiencing young-onset neurocognitive symptoms. In brief, BeYOND is a prospective observational cohort study evaluating clinical presentation, cognition, blood biomarkers and genetics. Recruitment criteria included individuals referred to the Neuropsychiatry Centre at the Royal Melbourne Hospital for a possible diagnosis of young-onset dementia who had psychiatric, behavioural, neurological and/or cognitive symptom onset prior to the age of 65 years. Participants were prospectively recruited during their index presentation. The study aimed to determine which combinations of clinical, cognitive, blood and genetic biomarker variables accurately differentiated causes of neurocognitive symptoms between the categories of early-onset AD (EOAD), non-AD neurodegeneration (nAD-ND) and PPD within this group (Supplemental File 1).

## Methods

### Participant recruitment

The BeYOND study protocol has been described previously, with participant recruitment from June 2019 to December 2020 and a follow-up over one year.(Loi, 2021) For this study, we evaluated cognition, genetics and blood biomarkers with clinical characteristics cross-sectionally. This project was approved by the Melbourne Health Human Research Ethics Committee (MH 2018.371).

### Diagnosis

Diagnosis was made according to consensus criteria including the McKhann criteria for AD(McKhann et al., 2011) and the Rascovsky criteria for behavioural-variant frontotemporal dementia.(Rascovsky et al., 2011) Cerebrospinal fluid protein levels of amyloid-β and phosphorylated-tau at threonine 181 (p-tau181) were utilized for diagnosis of AD. Psychiatric diagnoses were made according to the Diagnostic Statistical Manual, Fifth Edition.(American Psychiatric Association, 2013)

### Cognitive classification

Participants underwent formal neuropsychological assessments for the overarching domains of global cognition, memory (delayed recall) and executive function. Global cognitive function was determined using overall performance on the Neuropsychiatry Cognitive Assessment tool(Walterfang et al., 2006), which measures attention, memory, visuospatial, executive and language functions. A range of domain-specific neuropsychological tests were used to further assess memory and executive function, as shown in Supplementary Table 1.

Raw scores from the neuropsychological tests were converted into Z-scores based on normative data stratified by age and education as provided in the test manuals.(Griffith et al., 2022; Griffith et al., 2024) A Z-score was categorized into “impaired” at 1.5 standard deviations below the normative score or “normal” (not impaired”). If a participant completed more than one neuropsychological test in a particular domain, the classification for each test was determined. For differing classifications, two clinical neuropsychologists evaluated the data and formed a consensus opinion about the overall performance in that domain.

### Biomarker analyses

EDTA blood samples were collected from fasted participants and stored at −80°C. Plasma was tested in duplicates for selected biomarkers using Quanterix Simoa HD-X Neurology 2 Plex B for NfL and GFAP, and single Plex for p-tau181, according to the manufacturer’s specifications. ‘Age at assessment’ is the participant’s age at which blood samples were collected.

### APOE genotyping and polygenic risk score calculation

DNA was extracted from peripheral whole-blood samples and *APOE* genotype determined using TaqMan® genotyping assays (rs7412, assay ID: C____904973_10; rs429358, assay ID: C____3084793_20), (Life Technologies, Carlsbad, CA) on a QuantStudio 12K FlexTM Real-Time-PCR system (Applied Biosystems, Foster City, CA), using TaqMan® GTXpressTM Master Mix (Life Technologies) as per manufacturer instructions.(Porter et al., 2018a; Porter et al., 2018b) Allele loads were quantified for ε2 and ε4 and values were assigned to each individual (0,1 or 2) per allele, representing their *APOE* ε2 and ε4 status.

Participant genetic data were derived from an Axiom™ Precision Medicine Diversity Array (Applied Biosystems™) and imputed using the Haplotype Reference Consortium panel, for greatest cross-over of SNPs. Beta weights for PRS calculation and *p*-value thresholds for variant inclusion were sourced from the International Genomics of Alzheimer’s Project Alzheimer’s disease GWAS.^11^ PRS were calculated by first multiplying the number of effect alleles for each variant by the beta weights from the GWAS summary statistics, with the weighted variant scores summed for each individual. PRS were calculated at eight different GWAS *p-*value thresholds, from *p* < 5 x 10^-8^, reflecting the standard stringent GWAS-threshold, to *p* < 0.1, reflecting a suggestive-association threshold.

### Statistical analyses

R-package (version 4.3.1)(Team, 2023) was used for statistical analyses. Due to the potential bias that may arise with covariate-based imputations using relatively small sample sizes, missing values were not imputed. For non-parametric data, the Kruskal-Wallis test was used when comparing across three or more groups, with the Dunn test applied for post-hoc analyses. For non-parametric data comparison across two groups, the Wilcoxon rank sum exact test was used. For categorical data, Fisher’s exact test, with pairwise post-hoc analyses using the R package rcompanion(Mangiafico, 2023), was used. For other post-hoc analyses, Benjamini-Hochberg correction, with false discovery rate of 0.05 was used. *p_nom_* represent un-adjusted (nominal) *p*-values and *p_adj_* represent adjusted *p*-values using post-hoc analyses. Significance level α was set to 0.05.

For multinomial logistic regression, the R package nnet(Venables, 2002) was used. Only participants with complete data (on variables analysed) were used for modelling. Multinomial logistic regression was performed using diagnostic categories as the dependent variable and age at assessment, sex and individually-tested protein biomarker levels as the independent variables. *p_nom_*values were calculated using the Wald statistic.

The R package MuMIn(Bartoń, 2023) was used for information-theoretic model selection of general linear mixed models composed of all available participant variables. Only participants with complete data were analyzed. DeLong test was used to compare resulting models.

Plots were made the R packages ggplot2, ggbreak and ggstatsplot.(Patil, 2021)

## Results

### BeYOND participant demographics

Seventy-two participants were recruited, with 65 of them classified within the diagnostic categories of either EOAD (n = 18), nAD-ND (n = 23), or PPD (n = 24, Table 1). nAD-ND causes consisted of behavioural-variant frontotemporal dementia with definite frontotemporal lobar degeneration pathology (n = 6), probable behavioural-variant frontotemporal dementia (n = 4), Parkinson disease (n = 3), Niemann-Pick disease type C (n = 3), cerebellar degeneration (n = 2), cerebral autosomal dominant arteriopathy with subcortical infarcts and leukoencephalopathy (n = 1), progressive supranuclear palsy (n = 1), dementia with Lewy bodies (n = 1), vascular dementia (n = 1), and that of unclear etiology (n = 1). PPD causes consisted of depression (n = 7), schizophrenia (n = 4), bipolar affective disorder (n = 3), subjective cognitive impairment (n = 2), schizoaffective disorder (n = 2), obsessive compulsive disorder (n = 1), psychosis (n = 1), delusional disorder (n = 1), functional neurological disorder (n = 1) and ‘other specified mental disorder’ (n = 2). The EOAD, nAD-ND and PPD groups were similar for age of symptom onset, age at assessment, sex and *APOE* genotypes (Table 1). Missing data ranged between 0% and 26% (Table 1, Supplementary Table 2).

Of the seven individuals that were not classified as having one of the three diagnostic categories listed above, five had a diagnosis of mild cognitive impairment of unclear aetiology, one had autoimmune encephalitis and one had primary angiitis of the central nervous system. These seven individuals were excluded from further analyses as the primary aim of the study was to differentiate between the three aforementioned diagnostic categories.

### Cognitive profiles amongst diagnostic categories

Executive function was similar between the EOAD, nAD-ND and PPD diagnostic groups (*p_nom_* = 0.14) while global cognitive function and memory function were impaired in proportionally more EOAD than nAD-ND or PPD groups (Table 2 and Supplementary Figure 1). In particular, 100% of those with EOAD had global cognitive impairment, compared to 55% of those with PPD (*p_adj_* < 0.005). Similarly, memory was classified as impaired in 82% of those with EOAD, compared to 26% of those with nAD-ND and 46% of those with PPD (*p_adj_* < 0.005 for EOAD vs nAD-ND and *p_adj_*< 0.05 for EOAD vs PPD).

### Protein blood biomarker levels

Blood biomarkers levels for NfL, GFAP and p-tau181 were significantly different between the three categories, with levels higher in EOAD compared to the other categories (Figure 1, Table 2).

**Figure 1:**
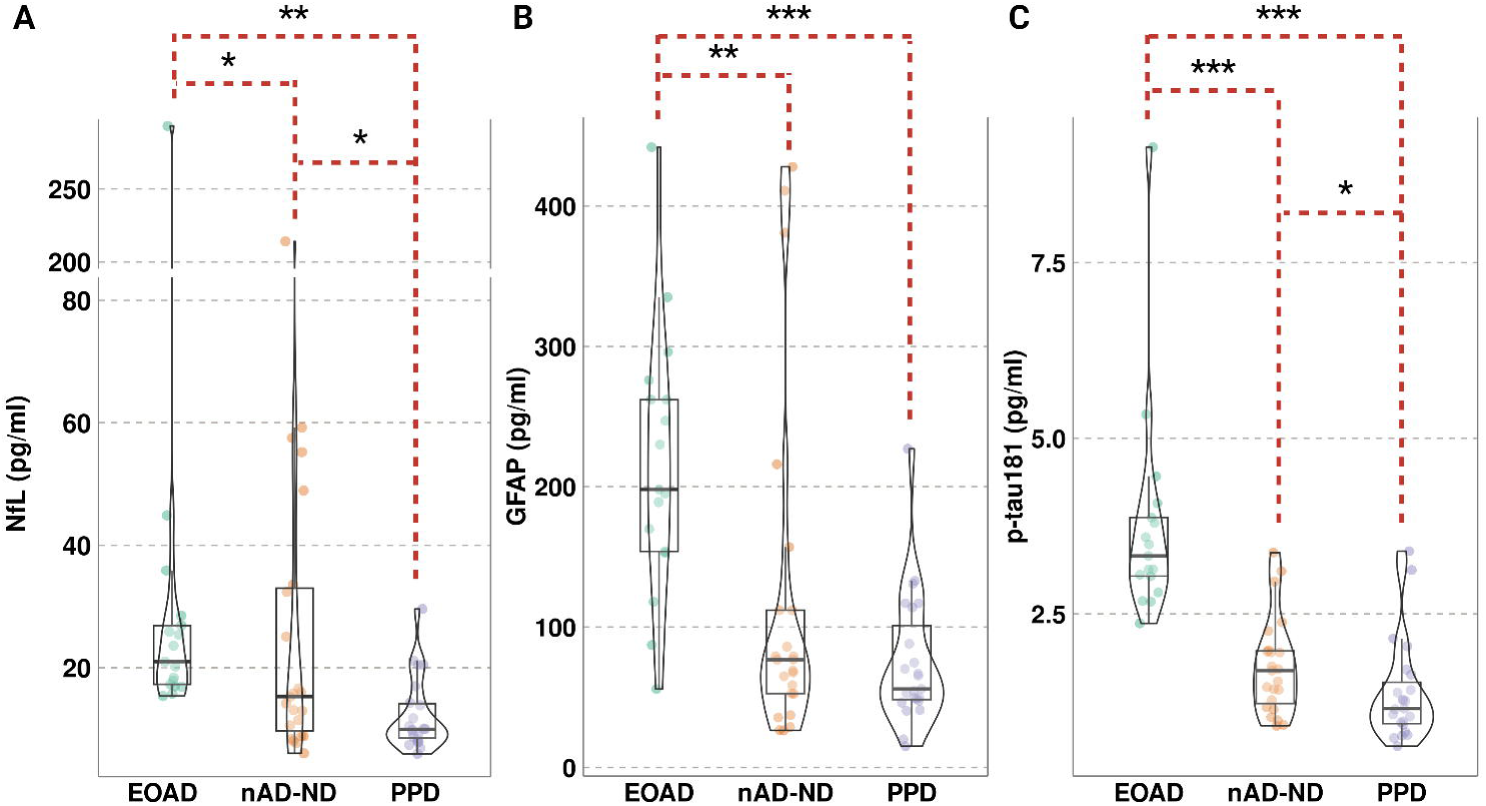
Blood biomarkers levels for NfL, GFAP and p-tau181 by diagnostic category. Violin plots of blood biomarker levels (pg/ml) for (A) neurofilament light chain (NfL), (B) glial fibrillary acidic protein (GFAP) and (C) phosphorylated-tau 181 (p-tau181) per diagnostic categories of early-onset Alzheimer’s disease (EOAD), non-AD neurodegeneration (nAD-ND) and primary psychiatric disorders (PPD). Boxplots represent the median and interquartile range. Adjusted (Dunn) *P*-value notation* *p_adj_* < 0.05, ** *p_adj_* < 5 x 10^-3^, *** *p_adj_* < 5 x 10^-5^.

Median NfL levels were 1.4-fold higher in participants with EOAD (21 pg/ml) compared to nAD-ND (15 pg/ml, *p_adj_*= 0.02, Supplementary Table 3) and 2.1-fold higher compared to those with PPD (10 pg/ml, *p_adj_* = 5.6 x 10^-5^). Median NfL levels were also elevated in nAD-ND compared to PPD (1.5-fold, *p_adj_*= 0.02).

In those with EOAD, median GFAP levels were 3.5-fold higher (198 pg/ml) than in those with PPD (56 pg/ml, *p_adj_*= 1.3 x 10^-5^) and nearly 2.6-fold higher those with nAD-ND (77 pg/ml, 1.8-fold, *p_adj_* = 4.0 x 10^-4^). While median GFAP levels were over 1.3-fold higher in nAD-ND compared to PPD, statistical significance was not found (*p_adj_* = 0.14).

Median p-tau181 levels were nearly 2-fold higher in EOAD (3.33 pg/ml) compared to nAD-ND (1.69 pg/ml, *p_adj_*= 3.2 x 10^-5^) and 2.9-fold higher compared to PPD (1.15 pg/ml, *p_adj_*= 1.9 x 10^-8^). Similarly, nAD-ND had 1.5-fold higher median p-tau181 levels than PPD (*p_adj_* = 0.04). Pairwise comparisons between diagnoses with the adjusted (Dunn) *p*-values for the three protein blood biomarkers are provided in Supplementary Table 3.

We further tested the association between protein blood biomarkers and diagnostic categories, adjusted for age at assessment and sex. Participants without missing data for these variables were analysed using multinomial logistic regression models. Demographic variables between the diagnostic categories of participants analyzed in these models were similar (Supplementary Table 4).

Compared to PPD, NfL demonstrated an odds ratio (OR) = 1.16 [95% CI: 1.05-1.29] for EOAD (*p_adj_* = 0.01) and OR = 1.16 [1.04-1.28] for nAD-ND (*p_adj_* = 0.01, Supplementary Table 5). Comparing between EOAD and nAD-ND, OR = 1.00 ([0.99-1.02], *p_adj_* = 0.61). While the OR for GFAP was statistically significant (*p_adj_* < 0.05) for EOAD vs PPD, the magnitude of the effect was quite small with OR = 1.02. Contrastingly, p-tau181 analyses yielded a large OR of 41.26 ([5.68-301.57], *p_adj_* = 0.002) and of 16.95 ([2.79-104.00], *p_adj_*= 0.006) for EOAD vs PPD and EOAD vs nAD-ND, respectively.

### Polygenic risk score associations

There were no statistically significant associations between PRS values, calculated using eight different LOAD GWAS^11^ *p-*value thresholds, and diagnostic categories (Supplementary Figure 2, Supplementary Tables 6 and 7). There was a trend for association using the stringent GWAS *p-*value threshold of 5 x 10^-8^ to calculate the PRS, with a median PRS value of 5.50 (interquartile range [IQR]: 4.14-6.14) in those with EOAD compared to 3.91 (IQR: 2.68-6.89) in those with nAD-ND and 2.45 (IQR: 0.70-4.69) in those with PPD (*p_nom_* = 0.085 for group comparison, Supplementary Figure 3).

### Information-theoretic model selection identifies key variables for pairwise discrimination between diagnostic categories

To determine which combinations of clinical and biomarker variables best discriminated between diagnostic categories, we utilized an information-theoretic approach for model selection. Models with the lowest Akaike information criterion with correction for finite sample sizes (AIC) were selected as they represented the optimal trade-off between model fit and model complexity.(Bartoń, 2023; Palmqvist et al., 2021) Variables included in the model selection were age at symptom onset, age at assessment, sex, cognitive function (global, memory and executive), blood biomarker levels (NfL, GFAP, and p-tau181), *APOE* ε2 and ε4 allele status and PRS (using GWAS *p*-value threshold of 5 x 10^-8^). Due to the model structure, only individuals with complete data could be analyzed, yielding 46 participants across the three diagnostic categories with similar demographic variables amongst them (Supplementary Tables 8-10).

For discriminating between EOAD and PPD within the BeYOND cohort, the best model (Model 1A, as denoted by producing the lowest AIC) contained five variables: *APOE* ε2 and ε4 status, global cognitive function, NfL and p-tau181 (Table 3). P-tau181 had the largest odds ratio of the included variables in Model 1, with an OR = 1.48 [95% CI: 1.33-1.65]. Interestingly, *APOE* ε2 and ε4 status both had OR < 1 for association with EOAD, compared to PPD. Three parsimonious models (Models 2A – 4A) equivalent to Model 1A (as denoted by having a ΔAIC < 2 compared to Model 1A) were identified. The area under the receiver operating curves (AUCs) for Models 1A-4A were very high (Figure 2A), ranging from 0.975 to 1.000 and were not statistically different (DeLong *p_nom_* > 0.05, Table 3). We next tested how models containing only one of the variables identified in Model 1A performed. Models 5A-7A and 9A were inferior to Models 1A-4A with respect to both ΔAIC and AUC values. Model 8A, containing only the p-tau181 variable, demonstrated a high AUC of 0.954 [95% CI: 0.887-1.000], but with a ΔAIC > 2, indicated an inferior model compared to Models 1A-4A. Assessing all models analysed for information-theoretic model selection revealed that p-tau181 had a sum of weights (SoW) = 1.00 (indicating this variable was present in all models), while only NfL, *APOE* ε2 and global cognitive function had a SoW ≥ 0.5 (indicating the importance of these variables as they were present in most of the well supported models based on AIC, Supplementary Figure 4).

**Figure 2:**
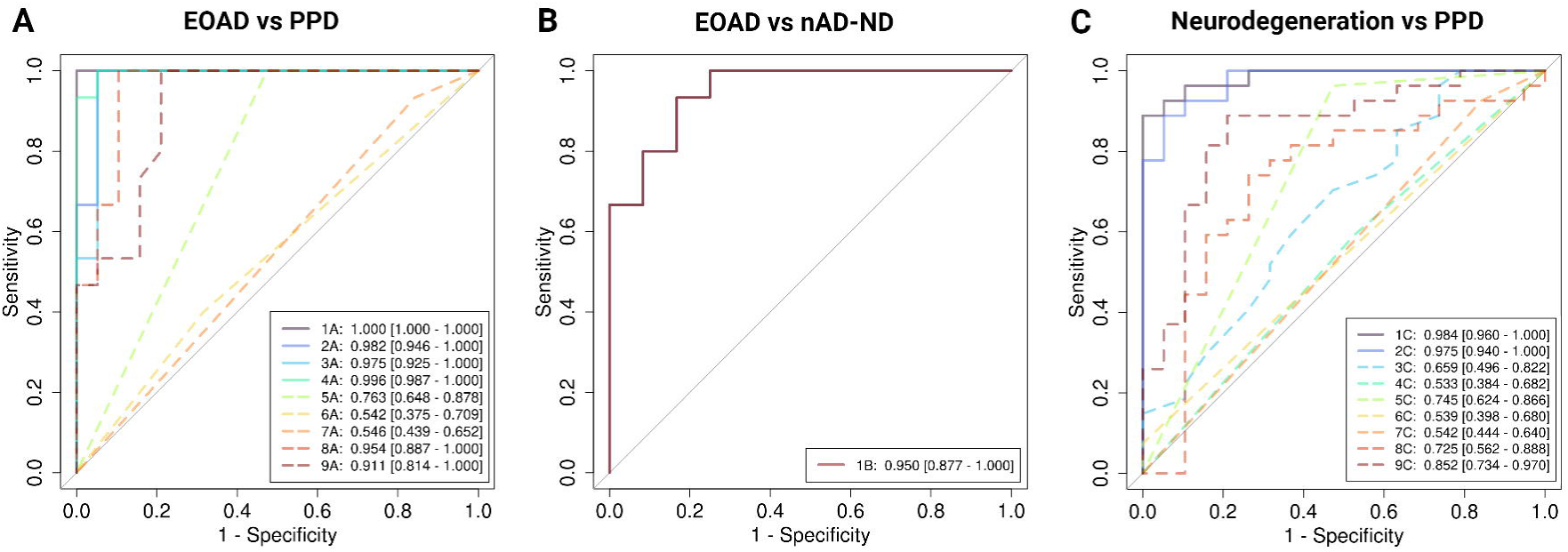
Area under the receiver operating curve plots for selected models discriminating between diagnostic categories. Best model (Model 1A/B/C), parsimonious models (solid lines beginning with Model 2A/B/C) and single variable models (dashed lines) in discriminating between A) early-onset Alzheimer’s disease (EOAD) and primary psychiatric disorders (PPD), B) EOAD and non-AD neurodegenerative cases (nAD-ND), and C) neurodegenerative cases (of all types) and PPD. Values for each model indicate area under the receiver operating curve (AUC) and 95% confidence interval. Variables included in each model for the different diagnostic category comparisons are listed in Table 3 and Supplementary Tables 11 and 12.

To discriminate EOAD from nAD-ND, the best model (Model 1B) required only the variable p-tau181 with an AUC of 0.950 [95% CI: 0.877-1.000] and an OR = 1.63 [95% CI: 1.38-1.93] (Supplementary Table 11, Figure 2B). P-tau181 had a SoW = 1 (Supplementary Figure 4). No other variable had a SoW ≥ 0.5, indicating that no other variables were considered significant for this comparison. In determining which variables would discriminate between all neurodegenerative cases (combining EOAD and nAD-ND) and PPD, the best model (Model 1C) contained seven variables: age at assessment, *APOE* ε2 and ε4 status, global cognitive function, PRS, p-tau181 and sex (Supplementary Table 12, Figure 2C), yielding an AUC = 0.984 [95% CI: 0.960-1.000]. Interestingly, global cognitive function had the largest magnitude of effect size with OR of 0.56 (95% CI: 0.43-0.71), followed by *APOE* ε2 (OR = 0.69), sex (OR = 1.37), and p-tau181 (OR = 1.28). One parsimonious models (Model 2C) contained six variables (PRS was not included) and demonstrated a similar AUC (0.975, 95% CI: 0.940-1.00, DeLong *p-*value = 0.35). Models 3C-9C, each containing only one variable from Model 1C, were all inferior to Models 1C and 2C with ΔAIC > 19. SoW for p-tau181 and global cognitive function was 1.00 and 0.99, respectively (Supplementary Figure 4). Other variables with SoW ≥ 0.5 included sex (0.91), age at assessment (0.73), *APOE* ε2 (0.71), PRS (0.58), *APOE* ε4 (0.52) and memory function (0.50), indicating that more variables may contribute to discriminating between neurodegeneration and PPD compared to between EOAD vs PPD and EOAD vs nAD-ND.

## Discussion

We determined how clinical, cognitive, protein blood biomarker and genetic variables differentiated causes of neurocognitive symptoms into the categories of EOAD, nAD-ND and PPD within the BeYOND cohort. We found that levels of NfL, GFAP and p-tau181 were higher in EOAD compared to nAD-ND and PPD. The association of the PRS with diagnostic categories did not reach statistical significance, but was suggestive. A model containing global cognitive function and levels of p-tau181 and NfL significantly discriminated between EOAD and PPD, while a model containing only p-tau181 significantly discriminated between EOAD and nAD-ND causes.

Blood biomarkers can differentiate and diagnose causes of late-onset cognitive impairment, especially LOAD.(Qu et al., 2021; Chouliaras et al., 2022; Hansson et al., 2023; Saunders et al., 2023) In particular, NfL has been implicated in a wide range of neurodegenerative conditions(Bacioglu et al., 2016) including LOAD(Mattsson et al., 2017), frontotemporal dementia(Rohrer et al., 2016) and amyotrophic lateral sclerosis.(Gaiani et al., 2017) This protein biomarker has shown utility in distinguishing neurodegenerative conditions from PPD.(Eratne et al., 2022; Eratne et al., 2024) In our cohort, we found that NfL could function similarly, with EOAD demonstrating higher blood levels than nAD-ND and PPD. NfL continued to differentiate between neurodegenerative and PPD cases when adjusted for age at assessment and sex, factors which can influence protein blood biomarker levels.(Simren et al., 2022; Tsiknia et al., 2022; Sarto et al., 2023; Eratne et al., 2024) The predictive power of NfL, however, was not demonstrated between EOAD and nAD-ND causes, supporting emerging data that NfL can be considered as a general marker of neurodegeneration.(Ashton et al., 2021)

We found elevated GFAP levels in EOAD compared to nAD-ND and PPD. However, the adjusted odds ratio for GFAP (OR = 1.01-1.02) was low for EOAD vs PPD and vs nAD-ND. This differs from a prospective study of 110 individuals with EOAD and 50 controls, where blood GFAP levels demonstrated an AUC of 96% in discriminating between these two groups.(Lv et al., 2023) A potential confounder in comparing these two studies is that the comparator group is individuals with PPD in our study, while healthy controls were used in the other, as well as the difference in samples sizes. Overall, our findings support the hypothesis that GFAP may be a marker in a number of neurodegenerative conditions.(Oeckl et al., 2019; Benussi et al., 2020; Cicognola et al., 2021; Abdelhak et al., 2022) More work is needed to elucidate what blood GFAP levels add in discerning the cause of neurocognitive symptoms, especially in the younger populations.

A strength of this study is the identification of specific variables that contribute to diagnosis, as the diagnosis of neurodegenerative diseases is moving towards a multi-omics approach.(Abdelhak et al., 2022) To the best of our knowledge, this is the first use of information-theoretic model selection(Bartoń, 2023) of multi-omic variables in a clinical cohort of young-onset neurocognitive disorders. A benefit of multi-model inference is the ability to determine important variables in a less biased manner as all studied variables can be included in these analyses, with the best model, as well as equivalent parsimonious models, identified. Using this approach, we found that global cognitive function and the blood biomarker levels of NfL and p-tau181 were sufficient to discriminate between EOAD and PPD. Similarly, p-tau181 alone was sufficient to discriminate EOAD from nAD-ND cases. Discriminating between all causes of neurodegeneration from PPD required more variables including *APOE* ε2 status, sex, p-tau181 levels, as well as global cognitive and memory function. These findings are similar to model selections performed in LOAD, where inclusion of p-tau181, *APOE* status, brain imaging and cognitive testing (memory and executive) were able to predict conversion to LOAD in individuals with subjective cognitive impairment or mild cognitive impairment.(Palmqvist et al., 2021) Additional studies of individuals with young-onset neurocognitive symptoms may reveal divergent variables between early-onset and late-onset causes. These findings support further hypothesis testing that single blood biomarkers may be used in the appropriate context to risk stratify individuals for neurodegeneration, which may be useful in both clinical care and clinical trials.(Hansson et al., 2022; Hansson et al., 2023; Self and Holtzman, 2023)

Multiple studies have demonstrated that p-tau181 is a very specific biomarker for LOAD.(Janelidze et al., 2020; Karikari et al., 2020; Thijssen et al., 2020) However, studies are lacking in describing the relationship between EOAD and p-tau181, especially in non-familial cases. One study did find elevated cerebrospinal fluid levels of p-tau181 in those with EOAD compared to healthy controls.(Kaur et al., 2020) Our study extends this finding to blood, with plasma p-tau181 levels demonstrating a robust ability to discriminate EOAD from other causes of young-onset neurocognitive symptoms.

AD-based PRS have been constructed using late-onset cases, which has shown strong ability to discriminate LOAD cases from healthy controls.(Bellenguez et al., 2022) In our cohort, there was a trend towards a higher PRS in EOAD compared to nAD-ND and PPD when restricting the PRS. Overall, the transferability of a PRS derived from a LOAD population to an EOAD population is mixed, with some studies demonstrating poor correlation.(Mantyh et al., 2023; Schott et al., 2016) Other studies have shown a potential role of a LOAD-derived PRS in predicting EOAD cases, though differences in age of participants and *APOE* status may confound direct comparisons.(Cruchaga et al., 2018; de Rojas et al., 2021; Fulton-Howard et al., 2021; Huq et al., 2021) These findings indicate the need for an EOAD-derived PRS, especially in individuals with non-familial cases.(Mol et al., 2022)

The relationship between *APOE* ε4 status and odds of being diagnosed with EOAD versus PPD was a surprising result, as the model selection suggested that a higher *APOE* ε4 allele count was associated with a higher probability of being diagnosed with PPD compared to EOAD. While *APOE* ε4 status is associated with a younger age-of-onset in LOAD and familial cases of AD, its role in sporadic cases of AD is less clear.(Olarte et al., 2006; Liu et al., 2013) This study was not powered to detail the relationship between *APOE* ε4 status, age of onset and diagnosis. Further work is needed to elucidate this relationship and to determine if our findings are due to differences in the genetic contributions to EOAD or limitations due to sample size.

There are additional limitations to our study that warrant careful consideration. Analysis based on the relatively small size of 65 participants from the BeYOND cohort must be acknowledged. Diagnostic heterogeneity, as to the specific cause of young-onset neurocognitive symptoms, was very high and necessitated formation of three diagnostic categories to perform analyses. The magnitudes of effect sizes greatly depend on the relative proportions of the different diagnostic categories within the study cohort. We attempted to mitigate this issue by accounting for covariates and performing information-theoretic model selection, reducing bias and penalizing model over-fitting. However, these are also affected, to a certain degree, by the diagnostic proportions included in each model. External cohort studies of young-onset neurocognitive symptoms are needed to validate our findings. With respect to biomarkers, we did not have access to standardized neuroimaging or emerging protein biomarkers like p-tau231, with the latter showing a differing role in LOAD diagnosis.(Self and Holtzman, 2023) We also performed PRS using a GWAS derived from LOAD cases;(Kunkle et al., 2019) there are recent LOAD-PRS with more implicated genetic loci.(de Rojas et al., 2021; Bellenguez et al., 2022) However, these recent studies use ‘proxy’ cases, where an individual who is asymptomatic is considered a GWAS case if they have at least one first degree relative diagnosed with LOAD, which may distort the relative contribution of GWAS loci to the risk of developing AD.(Escott-Price and Hardy, 2022) It is unclear how proxy cases affect LOAD GWAS transferability in EOAD cases.

The BeYOND cohort studied here is similar to other studies in its distribution of diagnostic causes for young-onset neurocognitive symptoms, thereby adding to this field of research and supporting the generalizability of our results.(Rossor et al., 2010; Loi et al., 2021) Overall, this study adds to our understanding of the multi-omic variables aiding in the diagnosis of young-onset neurocognitive symptoms. Our findings support further research into the use of protein blood biomarkers and cognitive profiles in the diagnostic pathway as well as identify the need for the development and validation of an EOAD PRS.

## Supporting information

Supplemental File - Tripod Checklist

## Data Availability

All data produced in the present study are available upon reasonable request to the authors.

## Acknowledgements

We would like to thank all the participants, their families and caregivers involved in the BeYOND study.

## Informed Consent Statement

All human subjects provided informed consent. This project was approved by the Melbourne Health Human Research Ethics Committee (MH 2018.371).

## Data Availability

Participant data access can be discussed with A/Prof Samantha M. Loi. Standard R packages were used for data analyses and can be sourced directly from public repositories.

