## Supplemental File - Tripod Checklist for "Blood biomarker profiles in young-onset neurocognitive disorders: a cohort study"

| **Section/Topic** | **Item** | **Checklist Item** | **Page** |
| --- | --- | --- | --- |
| **Title and abstract** | | | |
| Title | 1 | Identify the study as developing and/or validating a multivariable prediction model, the target population, and the outcome to be predicted. | 1 |
| Abstract | 2 | Provide a summary of objectives, study design, setting, participants, sample size, predictors, outcome, statistical analysis, results, and conclusions. | 2 |
| **Introduction** | | | |
| Background and objectives | 3a | Explain the medical context (including whether diagnostic or prognostic) and rationale for developing or validating the multivariable prediction model, including references to existing models. | 4-6 |
|  | 3b | Specify the objectives, including whether the study describes the development or validation of the model or both. | 4-6 |
| **Methods** | | | |
| Source of data | 4a | Describe the study design or source of data (e.g., randomized trial, cohort, or registry data), separately for the development and validation data sets, if applicable. | 6-8 |
|  | 4b | Specify the key study dates, including start of accrual; end of accrual; and, if applicable, end of follow-up. | 6 |
| Participants | 5a | Specify key elements of the study setting (e.g., primary care, secondary care, general population) including number and location of centres. | 6 |
|  | 5b | Describe eligibility criteria for participants. | 6 |
|  | 5c | Give details of treatments received, if relevant. | N/A |
| Outcome | 6a | Clearly define the outcome that is predicted by the prediction model, including how and when assessed. | 9-10 |
|  | 6b | Report any actions to blind assessment of the outcome to be predicted. | N/A |
| Predictors | 7a | Clearly define all predictors used in developing or validating the multivariable prediction model, including how and when they were measured. | 6-8 |
|  | 7b | Report any actions to blind assessment of predictors for the outcome and other predictors. | N/A |
| Sample size | 8 | Explain how the study size was arrived at. | N/A |
| Missing data | 9 | Describe how missing data were handled (e.g., complete-case analysis, single imputation, multiple imputation) with details of any imputation method. | 8 |
| Statistical analysis methods | 10a | Describe how predictors were handled in the analyses. | 9 |
|  | 10b | Specify type of model, all model-building procedures (including any predictor selection), and method for internal validation. | 8-9 |
|  | 10d | Specify all measures used to assess model performance and, if relevant, to compare multiple models. | 8-9 |
| Risk groups | 11 | Provide details on how risk groups were created, if done. | N/A |
| **Results** | | | |
| Participants | 13a | Describe the flow of participants through the study, including the number of participants with and without the outcome and, if applicable, a summary of the follow-up time. A diagram may be helpful. | N/A |
|  | 13b | Describe the characteristics of the participants (basic demographics, clinical features, available predictors), including the number of participants with missing data for predictors and outcome. | 9-10 |
| Model development | 14a | Specify the number of participants and outcome events in each analysis. | 9-10 |
|  | 14b | If done, report the unadjusted association between each candidate predictor and outcome. | 9-13 |
| Model specification | 15a | Present the full prediction model to allow predictions for individuals (i.e., all regression coefficients, and model intercept or baseline survival at a given time point). | N/A |
|  | 15b | Explain how to the use the prediction model. | N/A |
| Model performance | 16 | Report performance measures (with CIs) for the prediction model. | 13-15 |
| **Discussion** | | | |
| Limitations | 18 | Discuss any limitations of the study (such as nonrepresentative sample, few events per predictor, missing data). | 19 |
| Interpretation | 19b | Give an overall interpretation of the results, considering objectives, limitations, and results from similar studies, and other relevant evidence. | 15-20 |
| Implications | 20 | Discuss the potential clinical use of the model and implications for future research. | 19-20 |
| **Other information** | | | |
| Supplementary information | 21 | Provide information about the availability of supplementary resources, such as study protocol, Web calculator, and data sets. | 21 |
| Funding | 22 | Give the source of funding and the role of the funders for the present study. | 21 |

We recommend using the TRIPOD Checklist in conjunction with the TRIPOD Explanation and Elaboration document.
