## Supplemental Figures for "Blood biomarker profiles in young-onset neurocognitive disorders: a cohort study"

**Supplementary Figures**

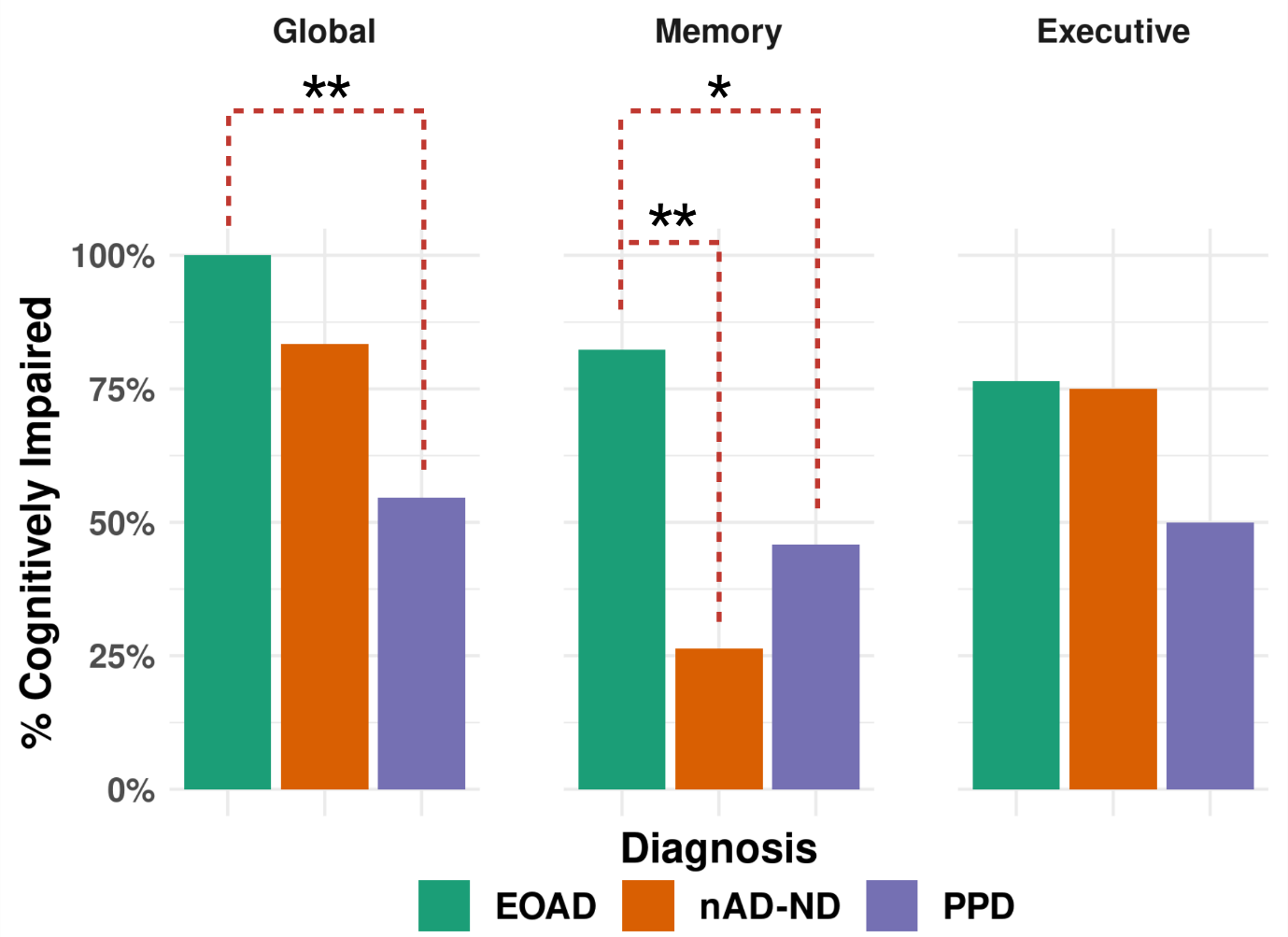

**Supplementary Figure 1:** **Comparison of impairment by cognitive domain and diagnostic categories.** Frequency of cognitive impairment for global cognitive, memory and executive function by diagnostic categories of early-onset Alzheimer’s disease (EOAD), non-AD neurodegeneration (nAD-ND) and primary psychiatric disorder (PPD). *, adjusted (Fisher) *p_adj_* < 0.05; **, *p_adj_* < 0.005.

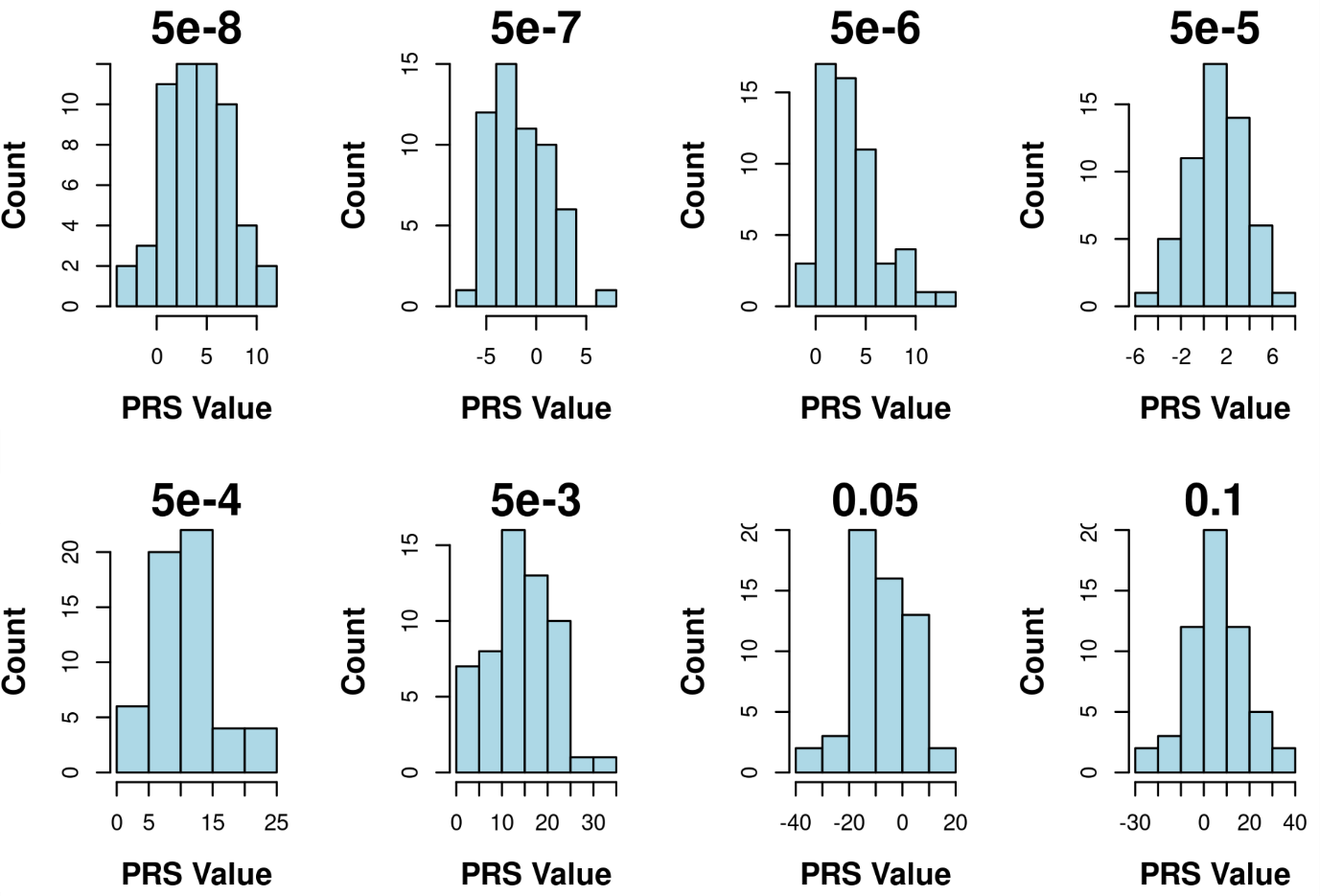

**Supplementary Figure 2: Distribution of polygenic risk score values by *p*-value thresholds.** Eight *p*-value thresholds were used to select single nucleotide polymorphisms identified by the Kunkel et al Alzheimer’s dementia genome wide association study(Kunkle et al., 2019) to calculate the polygenic risk score (PRS) for each participant. Frequency distributions are plotted for each *p*-value threshold (indicated by value above each plot).

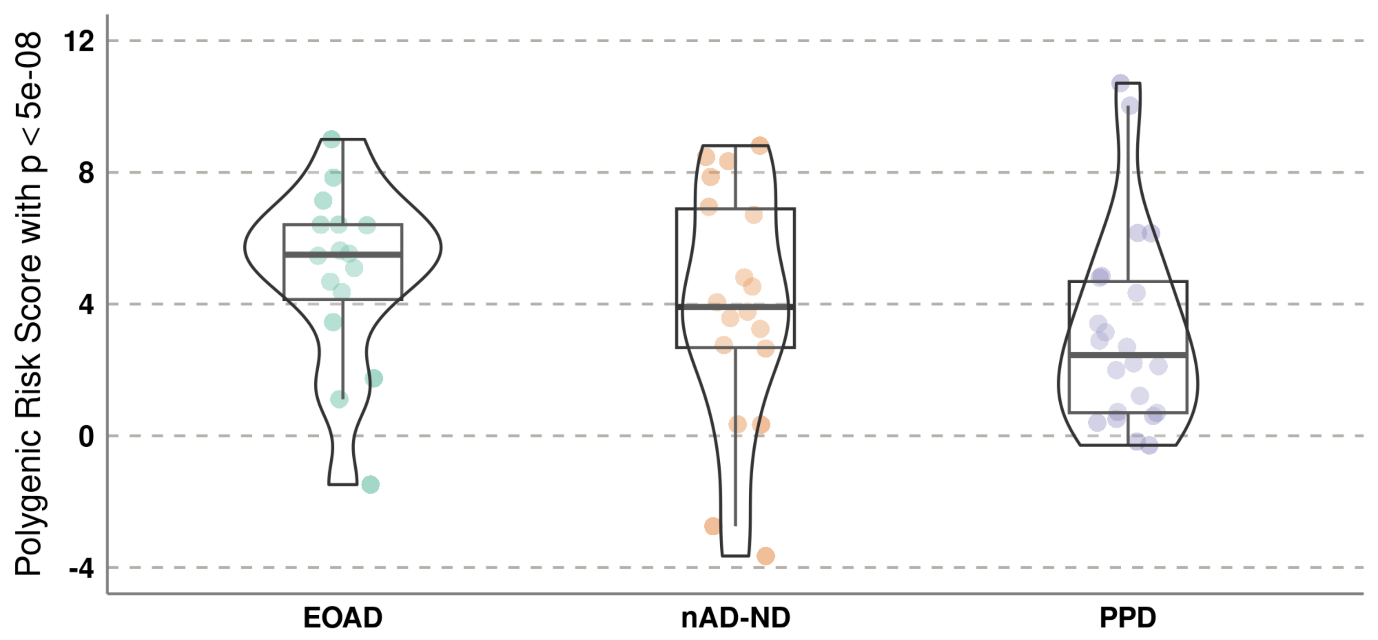

**Supplementary Figure 3: Distribution of polygenic risk scores with *p*-value threshold of 5 x 10^-8^ by diagnostic category.** Polygenic risk scores were calculated based on the Kunkel et al Alzheimer’s dementia genome wide association study(Kunkle et al., 2019) with *p*-value < 5 x 10^-8^ and scores were plotted for early-onset Alzheimer’s disease (EOAD), non-AD neurodegeneration (nAD-ND) and primary psychiatric disorders (PPD). Boxplots represent the median and interquartile range.

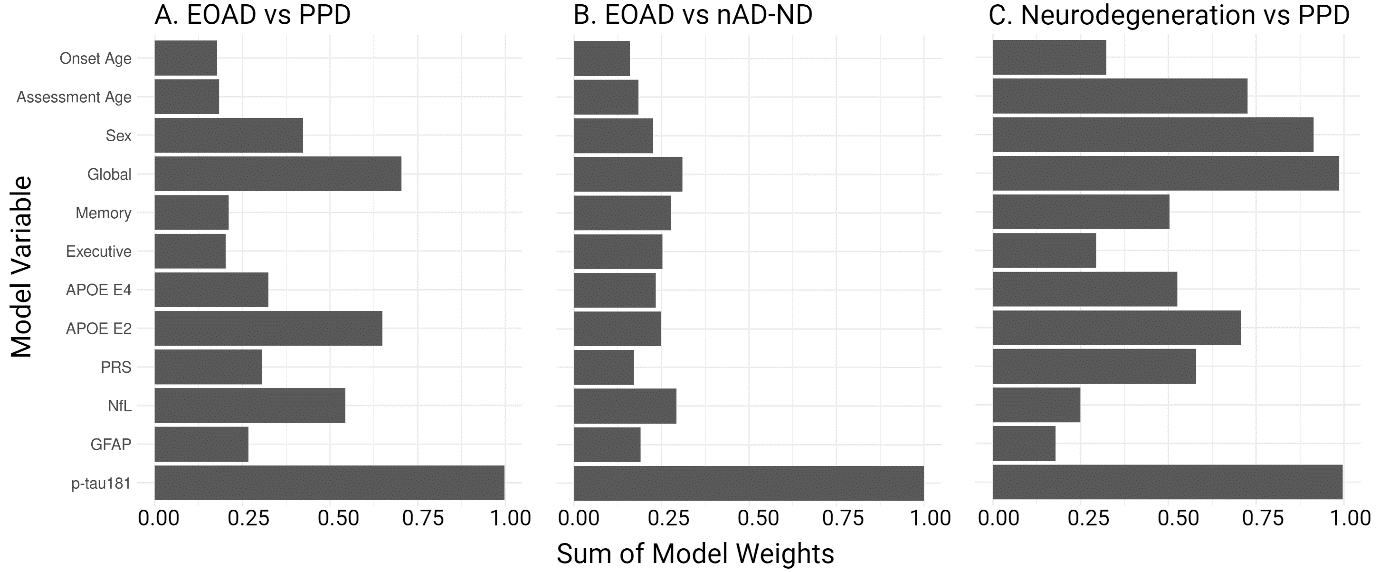

**Supplementary Figure 4: Sum of variable weights from information-theoretic model selection.** Sum of model weights (from all models tested during information-theoretic model selection for each variable, comparing A) early-onset Alzheimer’s disease (EOAD) and primary psychiatric disorders (PPD), B) EOAD and non-AD neurodegenerative cases (nAD-ND), and C) neurodegenerative cases (of all types) and PPD. PRS, polygenic risk scores calculated based on the Kunkel et al Alzheimer’s dementia genome wide association study (Kunkle et al., 2019) with *p*-value < 5 x 10^-8^.
